# Phase 1a Evaluation of LP-184 in Recurrent Glioblastoma: Safety, Pharmacokinetics, and Translational Optimization of CNS Exposure

**DOI:** 10.64898/2026.04.21.26351406

**Authors:** Karisa C. Schreck, Bachchu Lal, Jianli Zhou, Hernando Lopez-Bertoni, Matthias Holdhoff, Reginald Ewesuedo, Kishor Bhatia, Marc Chamberlain, John Laterra

**Affiliations:** Department of Neurology, Johns Hopkins University School of Medicine, Baltimore, Maryland; Sidney Kimmel Comprehensive Cancer Center, Johns Hopkins University School of Medicine, Baltimore, Maryland; Hugo W. Moser Research Institute at Kennedy Krieger, Baltimore, Maryland; Lantern Pharma Inc., Dallas, Texas

## Abstract

**Purpose:** Limited CNS bioavailability and pharmacodynamics are obstacles to effective systemic therapies for glioblastoma. One strategy to overcome these challenges is drug combinations enhancing CNS penetration and/or tumor chemosensitivity. LP-184, a synthetic acylfulvene class alkylator, induces DNA damage and inhibits glioblastoma cell viability in pre-clinical models. LP-184 is a prodrug converted to active metabolites by intracellular prostaglandin reductase 1 (PTGR1) that is over-expressed in >70% of glioblastoma. DNA damage induced by LP-184 is MGMT agnostic and reversed by transcription-dependent NER.

**Patients:** LP-184 was evaluated in a Phase 1a study (NCT05933265) in 63 adult patients with advanced malignancies including 16 patients with recurrent glioblastoma. All patients with glioblastoma received prior standard-of-care therapy and most had received 1 or more additional therapies before enrollment.

**Results:** Patients with glioblastoma experienced more frequent transaminitis, Grade 1-2 nausea and a trend towards more frequent and severe thrombocytopenia compared to the non-glioblastoma cohort. Otherwise, overall toxicity profiles were similar. Clinical pharmacokinetic analysis combined with published pre-clinical intra-tumoral bioavailability data (∼20% penetration) predicted that LP-184 at the recommended dose for expansion (RDE) would achieve cytotoxic levels if combined with spironolactone, a BBB permeable ERCC3 degrader and TC-NER inhibitor that sensitizes glioblastoma cells to LP-184 3-6-fold. We show that three daily doses of spironolactone deplete orthotopic glioblastoma PDX ERCC3 protein by ∼ 80% and increases tumor LP-184 cytotoxicity 2-fold.

**Conclusions:** LP-184 is well tolerated at the RDE, and we establish a clinically translatable scheme for dosing spironolactone in combination with LP-184 for a future Phase 1b clinical trial.

**Statement of translational relevance:** Treatment failure in glioblastoma reflects inadequate drug brain exposure and DNA repair– mediated resistance. LP-184, a novel acylfulvene alkylator, generates MGMT-independent DNA lesions predominantly repaired by transcription-coupled NER. In a Phase 1a dose finding trial, LP-184 was well-tolerated at the recommended dose for expansion (RDE) in participants with advanced cancers, including recurrent glioblastoma. Plasma drug levels achieved predicted effective systemic exposures but not brain concentrations based on projected 20% brain penetrance. Pharmacokinetic modeling indicates that NER inhibition could increase tumor chemosensitivity with the addition of spironolactone. The optimal dosing regimen for spironolactone combined with LP-184 was identified in orthotopic PDX models, facilitating advancement to Phase 1b/2a testing of LP-184 plus spironolactone.

## Introduction

Glioblastoma is the most common malignant primary brain tumor in adults and remains uniformly fatal despite maximal safe resection and radiotherapy with concurrent and adjuvant temozolomide (TMZ), with median overall survival of ∼15–18 months in protocol eligible patients (1, 2). Over half of patients with newly-diagnosed glioblastoma do not benefit from first-line chemotherapy with TMZ due to expression of O^6^-methylguanine-DNA methyltransferase (MGMT), which repairs temozolomide-induced DNA damage (3). Effective systemic options are limited at recurrence, and resistance driven by diffuse infiltration and marked intratumoral heterogeneity contribute to the substantial unmet need for novel therapies (4-6). An additional obstacle is the blood–brain barrier (BBB), which restricts central nervous system (CNS) exposure for most systemically administered drugs (7-9). CNS therapeutics face unique challenges in achieving sufficient intratumoral concentrations and targeting both in the enhancing and non-enhancing tumor compartments common in glioblastoma (10).

LP-184, an alkylator of the acylfulvene class, requires conversion of the parent prodrug to an active metabolite by intracellular oxidoreductases, particularly prostaglandin reductase 1 (PTGR1) (11). PTGR1 is overexpressed in many cancers relative to normal tissues, including glioblastoma, providing selective cytotoxicity for LP-184 (12-14). Importantly, the alkylating effect of LP-184 and other acylfulvenes is independent of MGMT status, with DNA repair primarily occurring by the transcription coupled nucleotide excision repair (TC-NER) or homologous recombination (HR) pathways (15-17). LP-184 demonstrates pre-clinical cytotoxicity as a single agent and anti-tumor activity is augmented in the setting of intrinsic and induced DNA repair deficiency and in combination with radiation (18-20).

Preclinical pharmacokinetic modeling and brain distribution studies further suggest that LP-184 crosses the BBB, with approximately 20% of systemic exposure reaching brain tumor and ∼10% reaching contralateral brain (20). Functional CNS delivery is supported by survival prolongation in orthotopic glioblastoma PDX models with good *in vivo* tolerability. Together, these data support clinical evaluation of LP-184 for glioblastoma, including assessment of safety, tolerability, and preliminary antitumor activity, alongside correlative analyses to better define CNS penetration and pharmacodynamic evidence of DNA damage in tumor and/or surrogate compartments.

In parallel, compounds that sensitize cancer cells to LP-184 were actively explored *in silico* and spironolactone was identified as a likely candidate. Spironolactone is a blood-brain barrier permeable aldosterone antagonist that inhibits DNA damage repair (specifically nucleotide excision repair; NER) by inducing proteasomal degradation of ERCC3 (XPB) (21-23). Preclinical studies demonstrate that spironolactone sensitizes glioblastoma models to LP-184 *in vitro* and *in vivo* as evidenced by increased tumor cell DNA damage response, increased tumor cell killing, and increased glioblastoma PDX response in vivo (20). By depleting ERCC3, spironolactone sensitizes glioblastoma cells to LP-184 resulting in a 3-6-fold lower IC_50_ in multiple glioblastoma cell models (24).

This study evaluates LP-184 in a first-in-human Phase 1a study in adult patients with advanced solid tumors to determine the recommended dose for expansion (RDE) (NCT05933265). Patients with recurrent glioblastoma were enrolled in this study, and here we report safety and tumor-specific outcomes in the glioblastoma cohort. We also identify a clinically translatable plan for dosing the ERCC3 degrader, spironolactone, in combination with LP-184 in preparation for a Phase 1b/2a study in patients with glioblastoma (NCT07431216).

## Methods

### Study design and treatment

NCT05933265 is a Phase 1a, first-in-human, dose escalation study evaluating the safety, tolerability, pharmacokinetics, and preliminary anti-tumor activities of LP-184 in adult patients (≥18 years old) with advanced solid tumors including glioblastoma. Key inclusion criteria included relapsed or refractory disease, Karnofsky performance status > 60, adequate organ function, measurable disease by RANO V2.0 (in glioblastoma patients), and life expectancy > 3 months. In glioblastoma patients, a chronic stable dose of dexamethasone was required as was prior initial treatment with surgery, temozolomide-based chemoradiotherapy and adjuvant temozolomide. There were no limits on the number of prior treatments.

Dose escalation followed a Bayesian optimal interval (BOIN) design with a starting dose level of 0.01 mg/kg. Dose limiting toxicities (DLT) were assessed during cycle 1. Patients received LP-184 IV over 30 (<u>⍰</u>5) minutes on days 1 and 8 of each 21-day cycle. Treatment continued until disease progression, unacceptable AEs, withdrawal of consent, loss to follow-up, protocol deviation, or at the discretion of the investigators.

### Endpoints and assessments

The primary objectives were to evaluate the safety and tolerability of LP-184 and determine the maximally tolerated dose (MTD) and recommended dose for expansion (RDE). Safety evaluations included clinical laboratory assessments, electrocardiograms, physical examination, vital signs, and adverse event (AE) monitoring. AEs were graded according to the NCI common terminology criteria for adverse events (CTCAE) version 5.0. The MTD was defined as the highest dose level at which the isotonic estimate of the dose limiting toxicity (DLT) rate is closest to the target toxicity rate of 0.3. The RDE was determined based on the totality of clinical data including pharmacokinetics, safety, and efficacy.

Secondary endpoints include pharmacokinetics and preliminary antitumor activities (i.e. objective response rate, duration of response and progression free survival). Additionally, archival tumor was analyzed for PGTR1 expression when available, however mandatory tissue submission was not required per protocol. Blood samples for pharmacokinetic analysis were collected at prespecified timepoints pre- and within 24 hours after the first infusion. A PK trough sample was taken on cycle 1 day 8. Plasma concentration of LP-184 was measured using a validated liquid chromatography-tandem mass spectrometry assay. Tumor assessment was performed at baseline, after every 2 cycles during the first 4 cycles, and every 4 cycles thereafter. Response was evaluated using RECIST V1.1 (non-glioblastoma patients) or RANO V2.0 (glioblastoma patients).

### PTGR1 quantification

Total RNA was extracted from sections of formalin-fixed, paraffin-embedded (FFPE) tissue blocks. Complementary DNA (cDNA) was synthesized from up to 2000 ng of RNA using the High-Capacity cDNA Reverse Transcription Kit (Thermo Fisher Scientific, part no. 4368814) according to the manufacturer’s instructions. Reverse transcription–quantitative polymerase chain reaction (RT-qPCR) was performed in triplicate using TaqMan probe–based assays for the target gene *PTGR1* (Thermo Fisher Scientific, part no. 4331182, assay ID Hs00400932_m1) and two housekeeping genes, *ACTB* (assay ID Hs01060665_g1) and *GAPDH* (assay ID Hs03929097_g1), with reactions prepared using a commercial master mix (Thermo Fisher Scientific, part no. 4369016) and run on a ViiA7 Real-Time PCR System under standard cycling conditions. Cycle threshold (Ct) values were obtained, and relative gene expression was calculated using the comparative ΔCt method, normalizing the target gene to the mean of the two housekeeping genes. No-template controls, a positive control (cDNA from a xenograft with high *PTGR1* expression), and a negative control (cDNA from a xenograft with low or undetectable *PTGR1* expression) were included in each run.

For determination of the preliminary PTGR1 threshold, RT-qPCR was performed as above on the following 10 cell lines: K562 (ATCC® CCL-243™), HCC-78 (ATCC® CRL-5872™), NCI-H1666 (ATCC® CRL-5885™), LNCaP (ATCC® CRL-1740™), U-87 MG (ATCC® HTB-14™), 8505C (ATCC® CRL-8305™), MDA PCa 2b (ATCC® CRL-2422™), LN-18 (ATCC® CRL-2610™), NCI-H1944 (ATCC® CRL-5907™), and NCI-H2228 (ATCC® CRL-5935™).

### Preclinical studies

#### Tumor Xenografts

The patient-derived glioblastoma xenograft line Mayo39 was obtained from the Mayo Clinic (Rochester, MN) (25) and cultured as previously described (24). For orthotopic xenografts, NOD-SCID mice received 10^5^ cells suspended in 2 µl PBS to the right caudate/putamen. To evaluate the kinetics of ERCC3 degradation, beginning on post-implantation day 15, mice received either 1, 2, or 3 doses of spironolactone (SP, 35 mg/kg i.p., q24 hours) or phosphate-buffered saline (PBS) control. Animals were sacrificed at the indicated times following spironolactone dosing and tissues isolated for immunoblot analysis. To determine how optimal spironolactone dosing effects tumor cell responses to LP-184, NOD-SCID mice were orthotopically implanted with Mayo39 cells as described above. On post-implantation day 15, mice began to receive 3 consecutive doses of spironolactone (35 mg/kg i.p., q24 hours) or PBS as control +/- LP-184 (4 mg/kg i.v.) administered 8 hours after the third dose of spironolactone. Animals were sacrificed 48 hours after LP-184 infusion and tissues processed for immunoblot.

#### Immunoblot analyses

Immunoblotting was performed as previously described (24) using NOVEX 4-12% Tris-glycine gradient gels (Thermo Scientific) and Amersham Protran nitrocellulose membranes (GE HealthCare). Membranes were probed with primary antibodies for ERCC3 (Aviva System Biology, ARP37963_P050), phospho-gammaH2AX (Cell Signaling, #2577S for immunoblot), cleaved caspase 3 (Cell Signaling #9664 for immunoblot) or actin (Cell Signaling cat #3700S) and secondary antibodies labeled with IRDye infrared dye (LI-COR Biosciences). Protein band intensities were quantified using the Odyssey Infrared Imager (LI-COR Biosciences) and analyzed using the Image Studio™ acquisition software from LI-COR imaging systems.

### Statistical analysis

Safety and efficacy analyses included all patients who received at least one dose of LP-184. Descriptive statistics were used to summarize safety events. Fisher’s two-sided exact test (95% confidence interval) was used to compare the frequency of AEs between the glioblastoma and non-glioblastoma group. Plasma concentration-time data from patients treated at dose 10 (n=11) were analyzed using nonlinear regression with a one-phase exponential decay model to estimate the dynamics of LP-184. For preclinical studies, One-way or Two-way ANOVA and Dunnett’s or Tukey post hoc tests were used when comparing multiple variables. Data analysis and visualization were conducted using R (version 4.1.0) and Graphpad PRISM (version 11.0.0).

## Results

### Patient characteristics and dosing

Sixteen patients with glioblastoma enrolled in the Phase 1a dose escalation study of LP-184 (63 patients were enrolled overall). See Table 1 for patient characteristics. Of these participants, all had glioblastoma, IDH-wild type, WHO grade 4. Eight of 14 with known MGMT status had an unmethylated promoter (57%). Molecular tumor profiling was completed on pre-treatment tissue from all tumors (Supplemental Table 1). Nine (56%) had a *TERT* promoter mutation, eight (50%) had *EGFR* amplification, five (31%) had homozygous deletion of *CDKN2A/CDKN2B*, five (31%) had the EGFRvIII translocation and three (18%) had *PTEN* single nucleotide variants.

**Table 1.** Glioblastoma patient characteristics.

| **Patients Characteristics** | **Total**<br>**(N=16)** |
| --- | --- |
| Age, years [median (range)] | 56 (22-72) |
| Sex, n (%) |  |
| Male | 10 (62%) |
| Female | 6 (38%) |
| Race, n (%) |  |
| White | 15 (94%) |
| Black | 1 (6%) |
| ECOG performance status, n/N (%) |  |
| 0 | 1/5 (20%) |
| 1 | 4/5 (80%) |
| Karnofsky performance status, n/N (%) |  |
| 70 | 3/11 (27%) |
| 80 | 1/11 (9%) |
| 90 | 7/11 (64%) |
| Number of Systematic Regimens, median (range) | 2 (1-4) |
| Prior Temozolomide, n (%) | 16 (100%) |
| Concurrent Temozolomide during RT, n (%) | 16 (100%) |
| Estimated Cycles of Temozolomide Maintenance post-RT, median (range) | 6 (0-15) |
| Temozolomide Rechallenge, n (%) | 2 (13%) |
| Temozolomide Discontinuation due to AEs, n (%) | 2 (13%) |
| Months between end of Temozolomide and start of LP-184, median (range) | 5 (1-44) |
| Prior Lomustine, n (%) | 3 (19%) |
| Number of Surgeries, n (%) |  |
| 1 | 13 (71%) |
| 2 | 3 (19%) |
| Number of Radiation Treatment, n (%) |  |
| 1 | 13 (71%) |
| 2 | 3 (19%) |
| Months between completion of radiotherapy and start of LP-184, median (range) | 11 (5-50) |
| IDH Status, n (%) |  |
| Mutated | 0 |
| Wildtype | 16 (100%) |
| MGMT Promoter Methylation, n/N with reported status (%) |  |
| Methylated | 6/14 (43%) |
| Unmethylated | 8/14 (57%) |
| Dexamethasone Dose at Enrollment, n (%) | 13 (81%) |
| 1 mg daily | 1 (6%) |
| 2 mg daily | 5 (31%) |
| 4 mg daily | 6 (38%) |
| 8 mg daily | 1 (6%) |

The study population was heavily pre-treated, with an average of two prior lines of therapy (range 1-5). All participants received prior radiation and temozolomide, with a median of 6 cycles of adjuvant temozolomide therapy. Subsequent therapies received at recurrence included lomustine (19%), re-resection (19%), or re-radiation (19%) and re-challenge with temozolomide (13%). The median time between prior adjuvant temozolomide and initiating study treatment with LP-184 was five months (range 1-44). The median time between completion of prior radiotherapy and initiating study treatment with LP-184 was eleven months (range 5-50). 88% of patients were on a stable dose of dexamethasone at time of study enrollment.

Patients with glioblastoma were enrolled at dose level 4 (DL4) and all but one subsequent dose levels (Supplemental Table 2). All patients with glioblastoma have come off study, with 14 (87.5%) patients having radiographic or clinical progression. The other two patients discontinued treatment for adverse events, one (DL8) having prolonged thrombocytopenia and neutropenia leading to dose holds, the other patient (DL12) had transient transaminitis and an elevated total bilirubin meeting Hy’s Law and the definition of a DLT.

### Toxicity

The range of treatment-related adverse events (TRAEs) in patients who received LP-184 on study with glioblastoma (n=16) was compared to participants with other solid cancers (n=47). The overall classes of toxicity were similar between the two cohorts, with no AEs observed exclusively in patients with glioblastoma (Figure 1a). Both nausea and alanine aminotransferase (ALT) increase were more common in patients with glioblastoma than other solid tumors (Fisher’s exact test, p<0.05). Nausea and/or vomiting was CTCAE Grade 1or 2 in all participants who experienced an AE. Of note, the study did not permit pretreatment with antiemetics during cycle 1 day 1; however prophylactic antiemetics were permitted for subsequent treatments. In patients with ALT elevation, one each Grade 3 and 4 AE was observed in patients with glioblastoma, while only Grade 1 ALT elevation was observed in other solid tumors. A nonsignificant trend toward increased rate and severity of thrombocytopenia also emerged in patients with glioblastoma (p=0.08).

**Figure 1.**
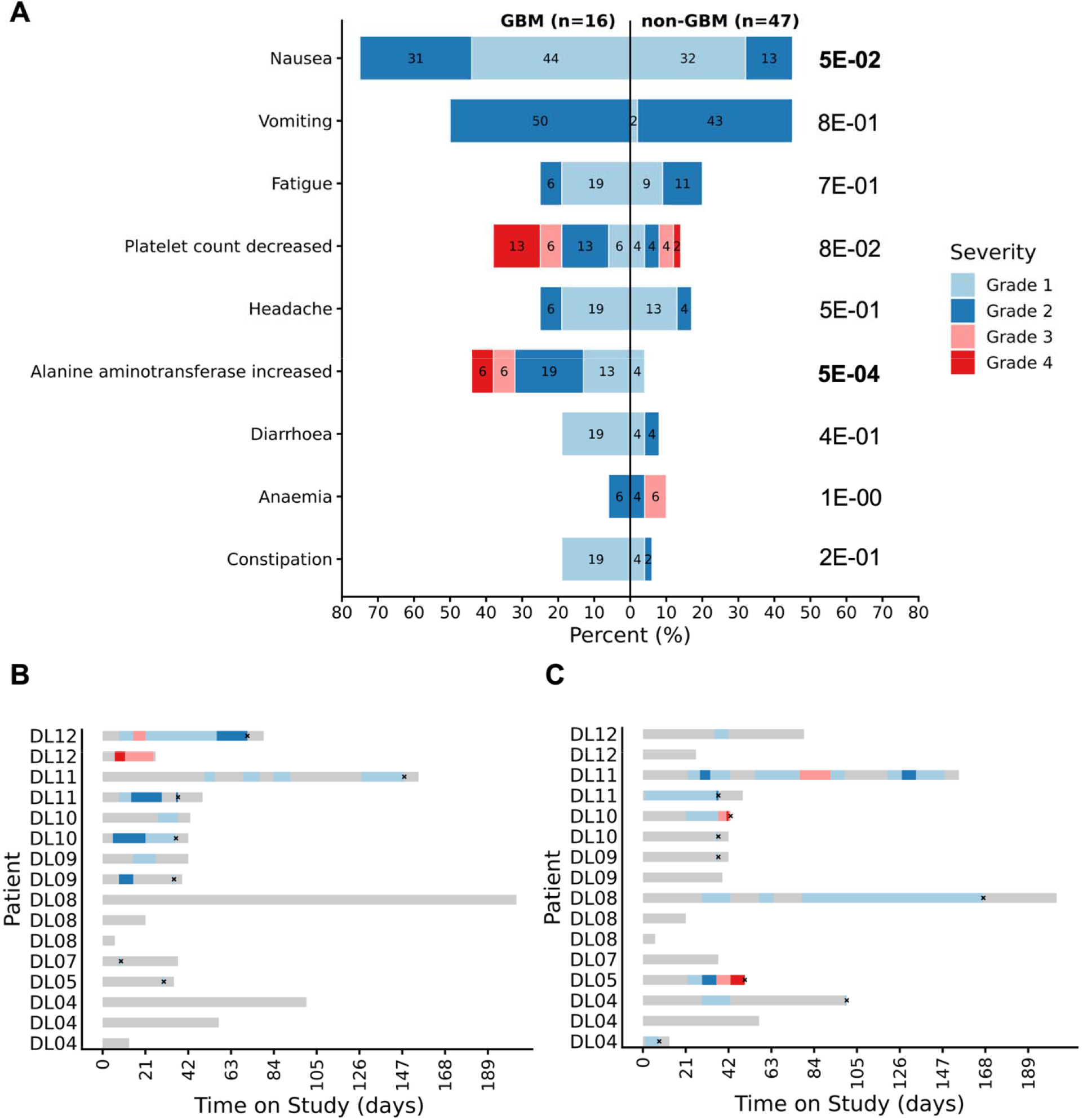
Glioblastoma patient safety profile (A) Common TRAEs (≥10%) in glioblastoma compared with non-glioblastoma patients; Two-sided Fisher’s exact test was conducted to test if AE proportions differ between two subgroups (p values were annotated on the side). (B) Swimmer plot of increased alanine aminotransferase based on reported lab values; (C) Swimmer plot of decreased platelet count based on reported lab values. Lack of reported follow-up lab values is denoted by x. GBM, glioblastoma, DL, dose level

We assessed whether the emergence of Grade 3-4 AEs was related to LP-184 dose. Both patients with Grade 3 or 4 ALT elevation received DL12, the highest dose (and established dose limiting toxicity) evaluated in the Phase 1a clinical trial (Figure 1b). No Grade 2 or higher ALT elevation occurred below DL9. Conversely, there was no clear trend between platelet count decrease and LP-184 dose level (Figure 1c).

We hypothesized that prior temozolomide exposure may have increased the risk of liver toxicity and thrombocytopenia given the known toxicity of temozolomide. However, there was no clear relationship between prior duration of temozolomide or the number of prior therapies and either ALT elevation or thrombocytopenia (Supplemental Figure 1).

### Preliminary responses

No radiographic responses were seen in patients with glioblastoma per RANO 2.0 (Figure 2). One patient (DL11) had stable disease for 4 months. A second patient (DL8) had radiographic progression on the first MRI response assessment, but continued treatment for clinical benefit per his treating physician’s discretion. This patient’s pathology (confirmed at re-resection prior to study entry) was revealed recurrent glioblastoma, IDH-wild type WHO grade 4, MGMT promoter methylated. The patient received a total of 6 months of LP-184 before discontinuing treatment for prolonged low-grade neutropenia and thrombocytopenia complicated by frequent dose holds. To date, the patient continues to have clinically and radiographically stable disease for more than 18 months after enrollment (>12 months off treatment per communication with treating physician) and has not received another line of therapy.

**Figure 2.**
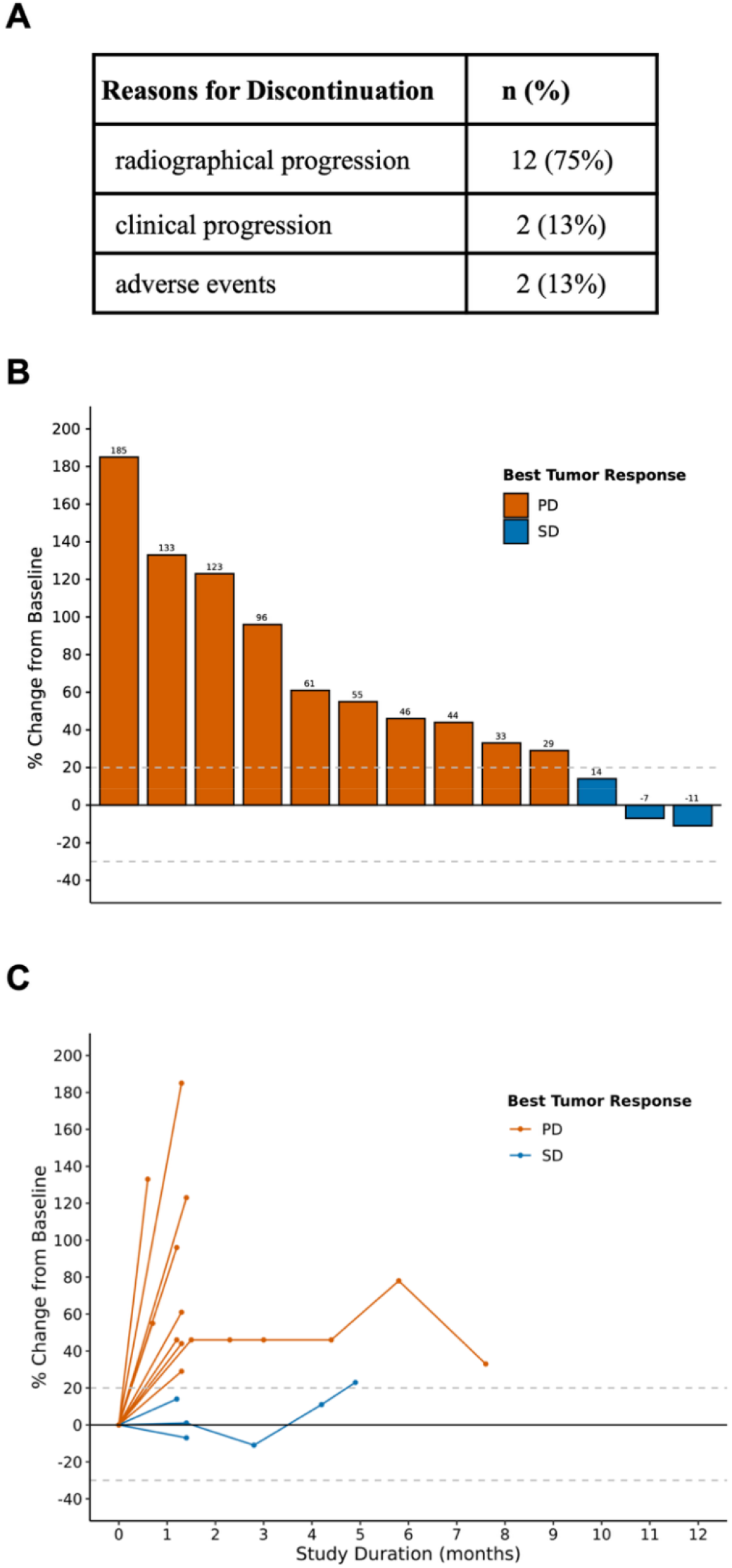
Glioblastoma patient response by RANO 2.0. (A) Reasons for treatment discontinuation; (B) Waterfall plot; (C) Spider plot. PD, progressive disease; SD, stable disease

### PTGR1 expression as a biomarker for sensitivity

Prostaglandin reductase 1 (PTGR1) facilitates conversion of the LP-184 prodrug to an active compound intracellularly. Because several attempts to quantify PTGR1 protein expression failed due to lack of reliable antibodies, RNA expression was used as a surrogate. IC50s from 78 previously screened cancer cell lines (26, 27) were plotted against PTGR1 RNA expression levels obtained from CellMiner CDB RNASeq data (28). A therapeutic LP-184 concentration of 300 nM was used as the sensitivity threshold, based on the average IC50 across solid tumor cell lines including human GBM isolates (288 nM) (20). This analysis identified a PTGR1 expression threshold of 4.5 log_2_(TPM+1) as the cutoff associated with *in vitro* LP-184 sensitivity (IC50 < 300 nM) (Figure 3a).

**Figure 3.**
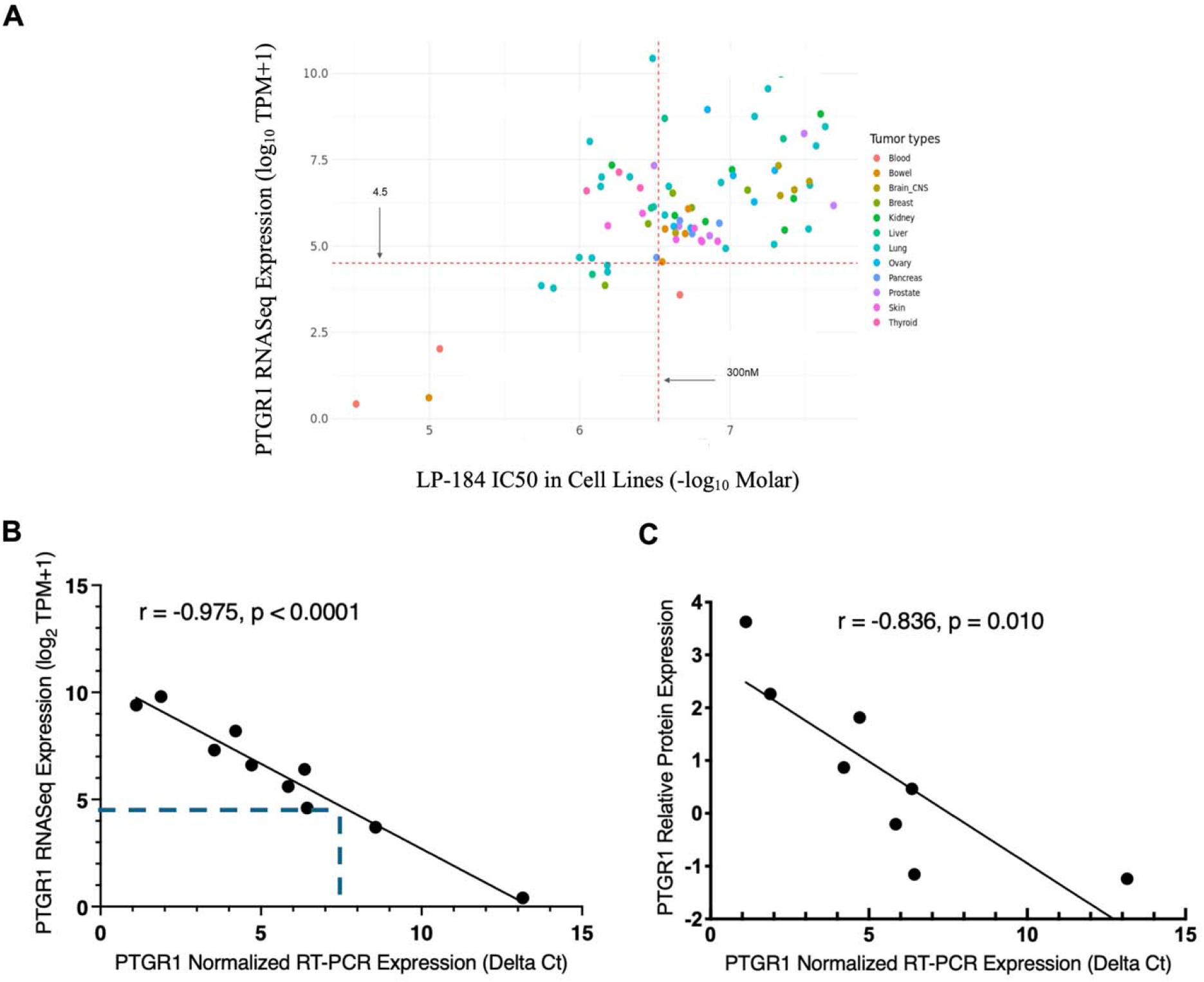
PTGR1 expression threshold measured by RT-qPCR. (A) IC50s from 78 previously screened cancer cell lines were plotted against PTGR1 RNA expression levels obtained from CellMiner CDB RNASeq data. PTGR1 expression level measured by RT-qPCR significantly correlated with the reported RNASeq expression (B) and relative PTGR1 protein expression (C). RNASeq, RNA sequencing; TPM, transcripts per million

To enable clinical quantification of *PTGR1*, an RT-qPCR assay was developed (see Methods), with expression normalized against two housekeeping genes (*GAPDH* and *ACTB*). Validation against CellMiner CDB data confirmed linear correlations between RT-qPCR and both RNASeq and protein expression across 10 cell lines. The 4.5 log_2_(TPM+1) RNASeq threshold corresponded to an RT-qPCR readout of approximately 7.5 ΔCT normalized PTGR1 (Figure 3b-c).

Pre-treatment tissue was available in 8/16 subjects with glioblastoma in the current Phase 1a study, and 75% of these samples demonstrated PTGR1 expression above the defined threshold.

### Pre-clinical assessment and modeling of spironolactone-induced ERCC3 degradation kinetics

With concerns that LP-184 administered at RDE (DL10) may not reach sufficient concentrations within the brain to have a tumoricidal effect as single agent, we sought to identify the translational potential of combining therapy with spironolactone. Spironolactone targets ERCC3 for ubiquination and proteasomal degradation and thereby sensitizes glioblastoma cells to LP-184 resulting in 3-6-fold lower IC50s (20). We evaluated how spironolactone dosing affects the timing and magnitude of ERCC3 protein degradation in an orthotopic glioblastoma PDX model to inform optimal spironolactone dosing for future clinical translation in combination with LP-184. Tumor ERCC3 levels were found to nadir at 8 hours (70% reduction) following a single dose of spironolactone (35 mg/kg i.p.) (Figure 4a). We used the 8-hour post-dose timing interval to examine the ERCC3 response to repetitive spironolactone dosing. Tumor ERCC3 levels were substantially lower (80% reduction) following three spironolactone doses (35 mg/kg i.p., q24 hour) compared to either one dose or two consecutive doses (49-57% reduction) (Figure 4b). Spironolactone had no effect on ERCC3 protein levels in liver (Figure 4c). Consistent with these results, the role of ERCC3 in DNA damage repair, and our prior published findings showing that spironolactone sensitizes GBM cells and orthotopic GBM PDXs to LP-184, we found that administering spironolactone (35 mg/kg i.p, q24 hours x3) prior to a single dose of LP-184 (4 mg/kg i.v.) increased the tumor cell death response (cleaved caspase 3) ∼ 2-fold compared to LP-184 alone (Figure 4d).

**Figure 4.**
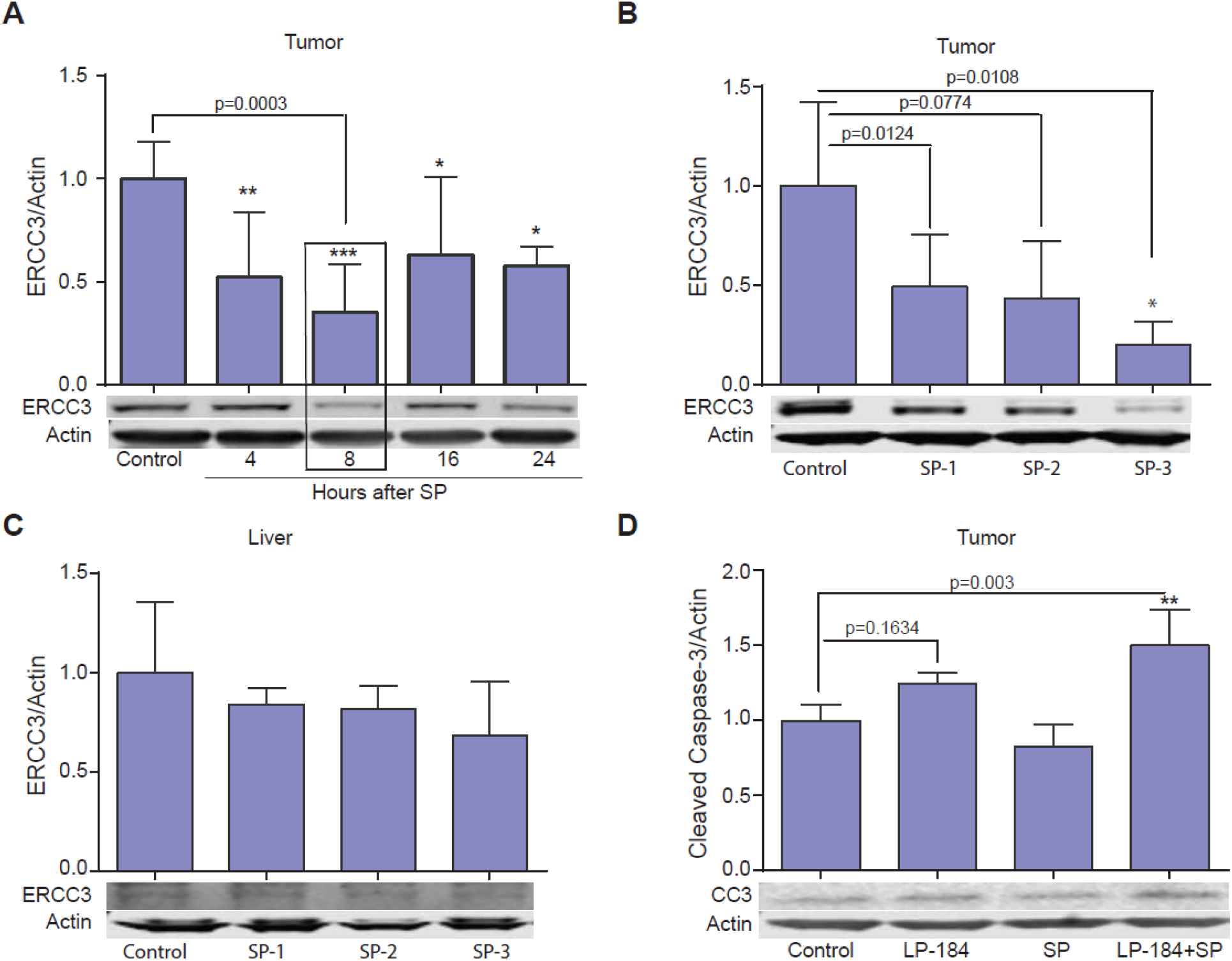
Spironolactone (SP) depletes ERCC3 in orthotopic GBM PDXs and enhances LP-184 tumor cytotoxicity. Mice (N=4) with pre-established orthotopic Mayo39 xenografts were treated with SP (35 mg/kg, i.p.) and/or LP-184 (4 mg/kg, i.v.) as described. Tissue-derived protein was analyzed by immunoblot, with representative blots shown. (A) Kinetics of tumor ERCC3 depletion following a single dose of SP. (B) Tumor ERCC3 levels quantified 8 hrs following one, two, or three doses of SP (SP-1, SP-2, SP-3, respectively) administered 24 hrs apart. (C) ERCC3 levels in livers obtained from mice treated as in panel B. (D) Tumor caspase-3 activation (cleaved caspase-3) in response to a single dose of LP-184 alone, SP alone (three daily doses as in panel B), or LP-184 administered 8 hrs following the last SP dose. Tumors were recovered for immunoblot analysis 48 hrs after LP-184 infusion.

### Pharmacokinetics

Plasma was collected at prespecified time points during and after study drug infusion from all patients participating in the Phase 1a study. Plasma concentrations of LP-184 reached 500nM (Cmax) in the lower dose cohorts of LP184 and were considerably higher (up to 900nM) at DL9-12 (Figure 5a-b; Supplemental Table 3). Notwithstanding, at all dose levels, plasma concentrations of LP-184 did not reach concentrations projected to achieve optimal on-target therapeutic concentrations in brain based on our preclinical findings of 20% tumor penetration and 10% normal brain penetration (in mice) and human GBM cell IC50s (50-300 nm) (20).

**Figure 5.**
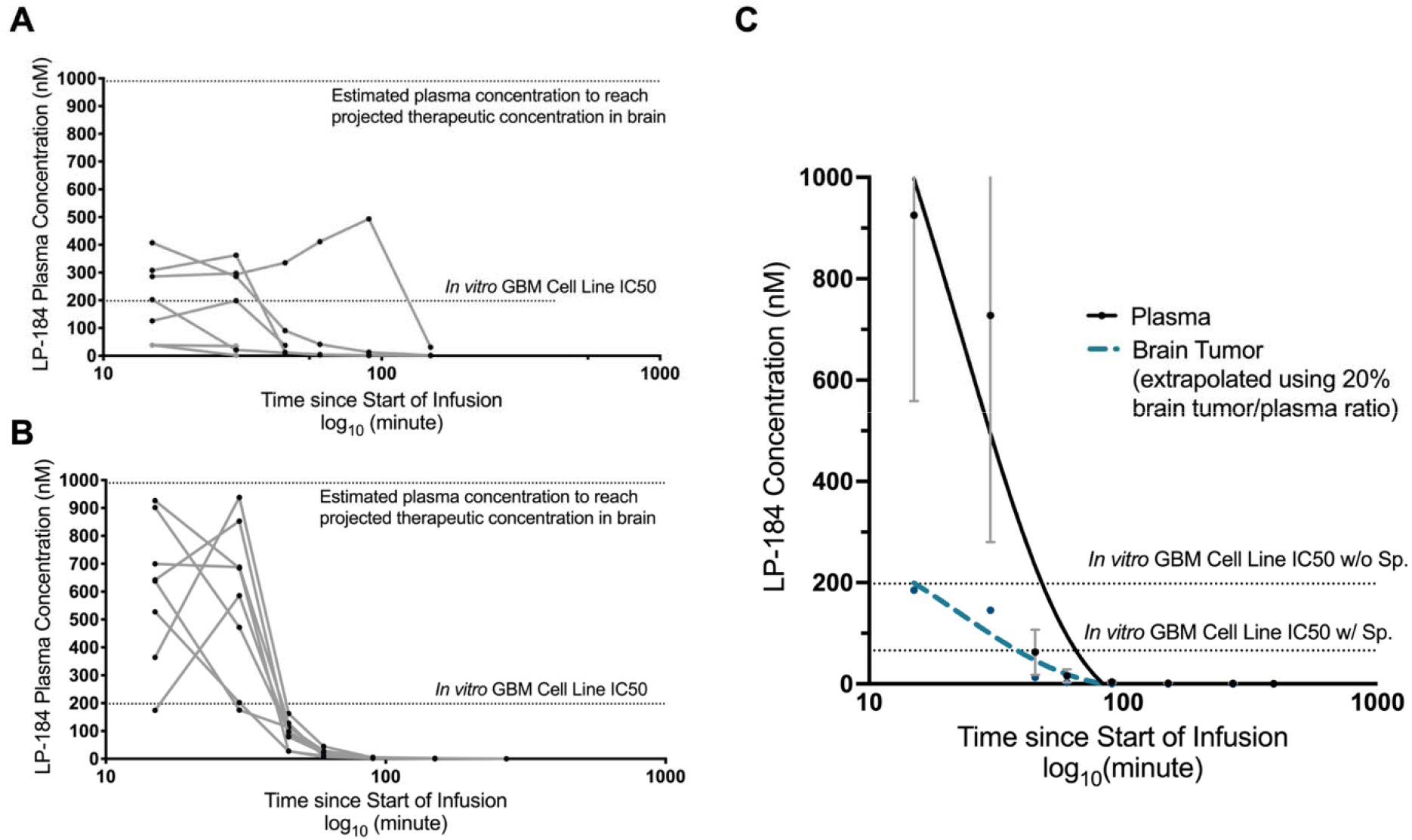
LP-184 pharmacokinetics (A) plasma concentration over time in glioblastoma patients DL04-DL08 (n=8); (B) plasma concentration over time DL09-DL12 (n=8); (C) Projected LP-184 therapeutic concentration in combination with Spironolactone (Sp.). Non-linear regression (on phase-decay) was used to generate LP-184 plasma concentration-time curves based on data from 11 patients treated at dose level 10 (0.39 mg/kg). Brain tumor concentrations were extrapolated using 20% brain tumor/plasma ratio. The IC50s with and without Sp. were derived from preclinical cell line data.

Using pharmacokinetic data from all patients treated with LP-184 at the recommended dose for expansion (DL10; n=11 including 2 with glioblastoma), we modeled projected LP-184 exposure in the presence of spironolactone (Figure 5c). These simulations predict that coadministration of spironolactone with LP-184 at the RDE (DL10, C_max_ geometric mean = 839 nM) will achieve intratumor LP-184 levels that exceed target cell IC50 ∼2-3 fold.

## Discussion

Alkylating agents are among the few chemotherapeutic classes incorporated into the standard of care for glioblastoma, and LP-184 exhibits properties that provide potential advantages compared to currently used agents. Temozolomide administered with radiation and following radiotherapy remains the standard for newly diagnosed glioblastoma (1). Temozolomide alkylates guanine, generating O6-methylguanine adducts that promote mispairing during replication and are processed by the mismatch repair pathway (MMR) (29). Lomustine, routinely used at recurrence, induces interstrand crosslinks that engage multiple DNA damage repair pathways, including Fanconi anemia (FA) and homologous recombination (HR) (30). The cytotoxic action of both temozolomide and lomustine are mitigated, at least in part, by MGMT activity, which removes methyl groups from guanine (3). In contrast, LP-184 preferentially alkylates adenine at the N3 position. The resultant DNA lesions are not susceptible to MGMT activity and predominantly resolved by transcription-coupled nucleotide excision repair (TC-NER) rather than base excision repair (BER) or global NER pathways (31). Like lomustine, cytotoxicity of LP-184 is enhanced in tumors with deficiencies in multiple DDR pathways, a vulnerability observed in EGFR-altered GBM based *in silico* analysis.

The potency of LP-184 in selected cancers can be enhanced via a machine learning-guided drug repurposing strategy. These analyses *in silico* identified EGFR alterations and low *ERCC3* expression as predictors of LP-184 sensitivity (20). Spironolactone, a diuretic and known inhibitor of TC-NER, degrades *ERCC3* and sensitizes tumors to LP-184 (20). Here, we show that daily spironolactone administration for 3 consecutive days before LP-184 substantially reduces ERCC3 protein in orthotopic GBM PDXs and increases tumor cell death compared to LP-184 alone. Moreover, based on the RDE pharmacokinetics, the LP-184-spironolactone combination is expected to exceed target brain concentrations in future clinical trial participants.

Patients with glioblastoma tolerated LP-184 supporting its potential for combination strategies and further study in patients with glioblastoma. LP-184 toxicity was broadly consistent with that observed in other solid cancers, although nausea, ALT elevation, and thrombocytopenia were more common in participants with glioblastoma. No clear association was observed between dose level and nausea or thrombocytopenia; in contrast, serious ALT elevation increased with dose. We hypothesized that prior temozolomide exposure might predispose to thrombocytopenia, given its known myelosuppressive effects, but did not detect a significant association between prior temozolomide and thrombocytopenia risk. This lack of association may reflect limited sample size or the relative uniformity of prior temozolomide exposure in the study glioblastoma cohort. Although no objective responses to LP-184 monotherapy were observed, one participant with a first recurrence of glioblastoma had early radiographic progression followed by stable disease for a total of eight cycles (6 months) of LP-184. The patient was taken off treatment for recurrent, low-grade myelosuppression resulting in frequent dose holds, but currently remains progression and treatment free more than 12 months after stopping LP-184 treatment.

LP-184 is a novel prodrug of the acylfulvene class that is well-tolerated and an identified RDE in solid cancers, including glioblastoma was determined. Its distinct alkylation site and activity independent of MGMT status support further development in glioblastoma and other malignancies characterized by deficiencies in DNA damage repair pathways (DDR). Nevertheless, its limited CNS penetration suggests that monotherapy is unlikely to achieve clinically meaningful activity in patients with brain cancers. Combination therapy with spironolactone, a BBB-permeable inhibitor of NER, shows promising preclinical efficacy. Here, we have identified the systemic concentration of LP-184 in human patients and the recommended dose for expansion for future clinical trials. Moreover, we have established a clinically translatable strategy for dosing the ERCC3 degrader, spironolactone, in combination with LP-184 in preparation for a Phase 1b/2a study in patients with first recurrence of glioblastoma following standard of care temozolomide based therapy (NCT07431216).

## Data Availability

All data produced in the present work are contained in the manuscript or available upon reasonable request to the authors.

## Acknowledgements

Funding for the clinical trial and preclinical studies included in this manuscript provided by Lantern Pharma Inc.

