## Supplemental tables and figure for "Phase 1a Evaluation of LP-184 in Recurrent Glioblastoma: Safety, Pharmacokinetics, and Translational Optimization of CNS Exposure"

**Supplemental material.**

**Supplemental Table 1.** Molecular features of study participants with glioblastoma. Gene alterations were retrieved from enrollment packages. PTGR1 RNA expression was assessed by RT-qPCR using archival tumor tissue as described in the method section.

| **Subject** | **IDH** | **MGMT**  **promoter methylation** | **EGFR alteration** | **PTEN mut** | **PI3K mut** | **TERT**  **promoter mut** | **ATRX mut** | **CDKN2 A/B deletion** | **PDGFR amplification** | **PTGR1 RNA Expression (ΔCt)** |
| --- | --- | --- | --- | --- | --- | --- | --- | --- | --- | --- |
| 1 | WT | Methylated | No | No | No | c.146C>T | No | CDKN2B del | No | NA |
| 2 | WT | Unmethylated | EGFR amp | No | No | c.146C>T | No | CDKN2A/2B codeletion | No | NA |
| 3 | WT | Unknown | No | p.R173C | No | c.146C>T | No | No | No | 5.97 |
| 4 | WT | Methylated | EGFRviii EGFR amp | ND | No | No | No | No | No | NA |
| 5 | WT | Unmethylated | EGFRviii EGFR amp | No | No | c.146C>T | No | CDKN2A/2B codeletion | No | 7.42 |
| 6 | WT | Unmethylated | EGFRviii EGFR amp | No | p.G376R | c.146C>T | No | No | No | NA |
| 7 | WT | Unmethylated | No | p.R335P | No | c.146C>T | No | No | No | NA |
| 8 | WT | Unmethylated | EGFR amp | No | No | No | No | No | No | NA |
| 9 | WT | Methylated | EGFR amp | No | No | No | No | No | No | 7.11 |
| 10 | WT | Methylated | EGFRviii EGFR amp | p.K128N | No | c.146C>T | No | No | No | NA |
| 11 | WT | Methylated | EGFRviii EGFR amp | No | p.P104R | c.146C>T | No | CDKN2A/2B codeletion | No | 7.09 |
| 12 | WT | Methylated | No | No | No | No | No | No | No | 9.18 |
| 13 | WT | Unmethylated | No | No | No | No | No | No | No | 7.32 |
| 14 | WT | Unmethylated | No | No | No | No | No | No | No | 7.88 |
| 15 | WT | Unmethylated | No | No | No | No | Mutant | No | No | 5.71 |
| 16 | WT | Unknown | No | No | No | c.146C>T | No | CDKN2B del | Amplified | NA |

**Supplemental Table 2.** Patient enrollment per dose level.

| **Treated per Dose Level** | **n (%)**  **Total N=16** |
| --- | --- |
| DL04 (0.07 mg/kg) | 3 (19%) |
| DL05 (0.11 mg/kg) | 1 (6%) |
| DL07 (0.20 mg/kg) | 1 (6%) |
| DL08 (0.25 mg/kg) | 3 (19%) |
| DL09 (0.31 mg/kg) | 2 (13%) |
| DL10 (0.39 mg/kg) | 2 (13%) |
| DL11 (0.49 mg/kg) | 2 (13%) |
| DL12 (0.61 mg/kg) | 2 (13%) |

**Supplemental Table 3.** LP-184 non-compartmental pharmacokinetic parameters derived from nominal plasma concentration-time data in glioblastoma patients.

|  | **DL04 (0.07 mg/kg)** | **DL05 (0.11 mg/kg)** | **DL7 (0.20 mg/kg)** | **DL8 (0.25 mg/kg)** | **DL9 (0.31 mg/kg)** | **DL10 (0.39 mg/kg)** | **DL11 (0.49 mg/kg)** | **DL12 (0.61 mg/kg)** |
| --- | --- | --- | --- | --- | --- | --- | --- | --- |
| **N** | 3 | 1 | 1 | 3 | 2 | 2 | 2 | 2 |
| **C_max_ (nM)** | 179 (3.8) | 199 (1.0) | 203 (1.0) | 179 (3.8) | 690 (1.5) | 738 (1.2) | 805 (1.2) | 741 (1.4) |
| **Auc_last_ (nM*hr)** | 99 (6.7) | 80 (1.0) | 54 (1.0) | 63 (6.1) | 255 (1.6) | 292 (1.4) | 365 (1.0) | 265 (1.5) |
| **T_max_ (min)** | 45 (15-90) | 30 | 15 | 20 (15-30) | 15 (15-15) | 23 (15-30) | 15 (15-15) | 30 (30-30) |
| **Half life (min)** | ND | ND | 31 | 15 (12-18) | 22 (19-24) | 37 (17-56) | 18 (12-23) | 18 (14-21) |

C_max_, and AUC_last_, are reported as geometric mean (geometric standard deviation factor); Half-life (apparent elimination half-life) and T_max_ are reported as mean (minimum-maximum); C_max_, maximum plasma concentration; AUC_last_, area under the curve to the last quantifiable plasma concentration; T_max_, time required to reach the maximum plasma concentration;  ND, not determined

**Supplemental Fig. 1.** Association between number of prior treatments or prior TMZ duration with increased ALT (A, B) and decreased PLT (C, D). TMZ, temozolomide; ALT, alanine aminotransferase; PLT, platelet count.

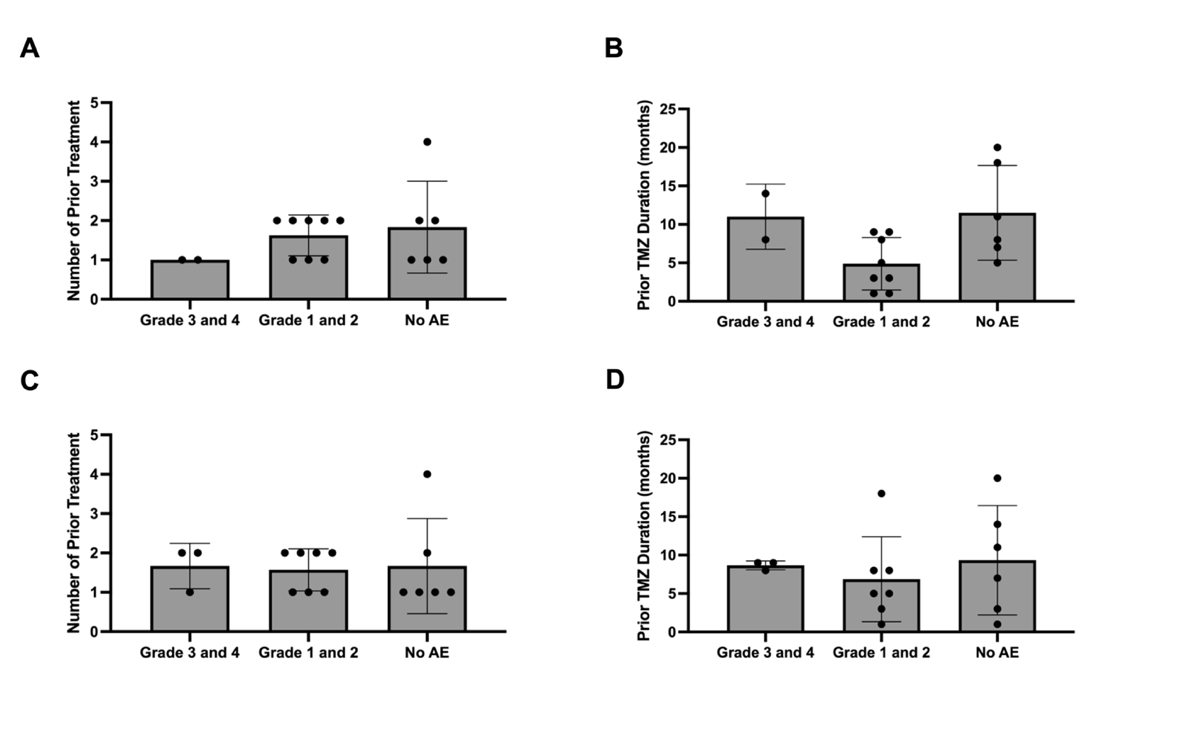
